# Predictors for Reactogenicity and Humoral Immunity to SARS-CoV-2 Following Infection and mRNA Vaccination: A Regularized Mixed-Effects Modelling Approach

**DOI:** 10.1101/2022.04.05.22273450

**Authors:** Erin C. Williams, Alexander Kizhner, Valerie S. Stark, Aria Nawab, Daniel D. Muniz, Felipe Echeverri Tribin, Juan Manuel Carreño, Dominika Bielak, Gagandeep Singh, Michael E. Hoffer, Florian Krammer, Suresh Pallikkuth, Savita Pahwa

**Affiliations:** Department of Otolaryngology, University of Miami Miller School of Medicine, Miami, Florida, 33136 USA; Department of Microbiology and Immunology, University of Miami Miller School of Medicine, Miami, Florida, 33146, USA; Department of Biomedical Engineering, University of Miami, Miami, Florida, 33136, USA; University of Miami Miller School of Medicine, Miami, Florida, 33136, USA; Department of Neurological Surgery, University of Miami, Miller School of Medicine, Miami, Florida, 33136, USA; Department of Microbiology, Icahn School of Medicine at Mount Sinai, New York, New York, 10029, USA; Department of Pathology, Molecular and Cell-based Medicine, Icahn School of Medicine at Mount Sinai, New York, New York, 10029, USA

**Keywords:** COVID-19, protective antibodies, vaccine reactogenicity

## Abstract

**Background:** The influence of pre-existing humoral immunity, inter-individual demographic factors, and vaccine-associated reactogenicity on immunogenicity following COVID vaccination remains poorly understood.

**Methods:** Ten-fold cross-validated least absolute shrinkage and selection operator (LASSO) and linear mixed effects models were used to evaluate symptoms experienced during natural infection and following SARS-CoV-2 mRNA vaccination along with demographics as predictors for antibody (AB) responses in COVID+ participants in a longitudinal cohort study.

**Results:** In previously infected individuals, AB were more durable and robust following vaccination when compared to natural infection alone. Higher AB were associated with experiencing dyspnea during natural infection, as was the total number of symptoms reported during the COVID-19 disease course. Both local and systemic symptoms following 1^st^ and 2^nd^ dose of SARS-CoV-2 mRNA vaccines were predictive of higher AB after vaccination, as were the demographic factors of age and Hispanic ethnicity. Lastly, there was a significant temporal relationship between AB and days since infection or vaccination.

**Conclusion:** Vaccination in COVID+ individuals ensures a more robust immune response. Experiencing systemic and local symptoms post-vaccine is suggestive of higher AB, which may confer greater protection. Age and Hispanic ethnicity are predictive of higher AB.

## Introduction

The heterogeneous presentation of severe acute respiratory syndrome coronavirus 2 (SARS-CoV-2) is associated with inter-individual factors [1, 2], including age, biological sex, comorbidities, individual susceptibility to the virus, exposure load, viral shedding, pre-existing binding or neutralizing antibodies [3, 4], and pre-existing cross-reactive T cells [5-7]. Variability in these factors and their distinct contributions to the individual immune response has made it difficult to generalize the clinical disease course of SARS-CoV-2-infected individuals [1, 8]. Immunoassays have been used extensively throughout the COVID-19 pandemic [9]. Their primary utility lies in characterizing the immune response following vaccination, assessing viability for convalescent plasma donation, and acting as a population surveillance tool [10, 11] though the most pressing work remains developing correlates for protection. Neutralizing and binding titers remain well supported as protective markers [3, 12] regardless of natural infection or vaccination, including a recent study [13] which associated increased binding and neutralizing antibodies with an inverse risk for SARS-CoV-2 infection following mRNA-1273 (Moderna) vaccination.

Previous studies have evaluated change in peak post-vaccination antibody titers as a function of time [14] and the relationship between quantitative antibodies and disease severity [15]. Additionally, there is evidence suggesting that antibody titers in vaccinated, previously coronavirus disease 2019 (COVID-19) positive individuals [16-19] can predict the degree of individual immune protection [20]. The aim of this paper is to investigate relationships between sociodemographic factors, reactogenicity, and immunogenicity following SARS-CoV-2 mRNA vaccination in previously SARS-CoV-2 infected participants. This analysis addresses a gap in the current COVID-19 literature that may help elucidate how pre-existing immunity and vaccine reactogenicity are associated with post-vaccination antibody titers (i.e., humoral immunogenicity).

While a reactogenic response (i.e., short-term symptoms such as injection-site pain, fever, myalgia, etc.) is expected following vaccination [19], there is no current evidence to support a relationship between reactogenicity and immunogenicity for COVID-19 vaccines. We also address a major clinical question regarding how severity of the COVID-19 disease course influences immunogenicity following SARS-CoV-2 vaccination. Conclusions drawn from this study may contribute to a more personalized public health approach to future COVID-19 vaccine strategies, which could account for an individual’s demographics (e.g., age, gender, or race) or existing immunity prior to vaccination receipt [21].

## Methods

### Study Design and Participants

We included a subset of individuals who are enrolled in our IRB-approved (#20201026), longitudinal, prospective SARS-CoV-2 immunity study. Following the written informed consent process, participants answered questions detailing their demographics, lifestyle habits, past medical history (including COVID-19), and COVID-19 infection symptoms. Blood samples were collected for serum and peripheral blood mononuclear cell (PBMC) processing. Plasma was stored at -80°C and PBMCs were cryopreserved in liquid N2 [22]. All participants agreed to sample banking for future research use.

Participants who were enrolled in the study between October 2020 – June 2021 and had a history of COVID-19 were included in this analysis. mRNA vaccines (Pfizer (BNT162b2) and Moderna (mRNA-1273)) were the only options available [9] during the enrollment period and therefore most participants received mRNA vaccines. Individuals who only received one dose of an mRNA vaccine or received the Johnson & Johnson vaccine were excluded. Additionally, participants who were administered their second dose of an mRNA vaccine >7 days after or <4 days before the recommended [23] number of days after the first dose (21 days for Pfizer; 28 days for Moderna) were excluded in order to account for the temporal, transient nature of post-vaccine reactogenicity and the subsequent immune response in order to best reflect the general population. Individuals with SARS-CoV-2 re-infection or breakthrough infection were also excluded. All samples provided during the baseline visit and thereafter were included in this analysis.

Participants who received vaccines were instructed to return for two additional visits, where they answered binary “Yes/No” questions in a survey about their symptoms following vaccination. Symptoms were rated on a Likert scale, where a “0” indicated no symptoms and a “10” indicated the highest symptom severity. They also provided blood samples, which were processed for PBMCs and plasma as described above. Fourteen days after participants received Dose 2, we classified them as “fully vaccinated”.

### Enzyme-Linked Immunosorbent Assay (ELISA)

SARS-CoV-2 ELISAs were performed using a well-described assay developed by the Icahn School of Medicine at Mount Sinai [10, 11]. Briefly, 96-well plates were coated at 4 °C with SARS-CoV-2 spike protein (2 μg/ml) solution and incubated overnight. Plates were blocked with 3% non-fat milk prepared in PBS with 0.1% Tween 20 (PBST) and incubated at room temperature for 1h. After blocking, serial dilutions of heat inactivated serum samples were added to the plates and incubated for 2h at room temperature. Plates were washed three times with 0.1% PBST followed by addition of a 1:3,000 dilution of goat anti-human IgG–horseradish peroxidase (HRP) conjugated secondary antibody (50μl) well and incubated 1h. Plates were washed, 100 μl SIGMAFAST OPD (o-phenylenediamine dihydrochloride;) solution was added to each well for 10 min and then the reaction was stopped by the addition of 50μl per well of 3M hydrochloric acid. The optical density at 490nm (OD490) was measured using a Synergy 4 (BioTek) plate reader. The background value was set at an OD490 of 0.15 then discrete titers were reported in values of 1:100, 1:200, 1:400, 1:800, 1:1600, 1:3200, 1:6400, 1:12800, 1:25600, 1:51200, 1:102400, and 1:204800. The limit of detection was set at 1:100.

### Statistical Analysis

SARS-CoV-2 antibody titers were log2-transformed before all statistical analyses. We utilized a generalized additive model to model the bi-phasic change in antibodies over time, including the rate of antibody decay following natural infection (after last positive SARS-CoV-2 test [LPT] result) and full vaccination (≥14 days after the second mRNA vaccine dose) [24] by modelling antibody titers with the smooth function number of days elapsed using a cubic regression with 3 knots and the fixed effect of vaccination status (after natural infection/pre-vaccination and after full vaccination). We replicated the above as a linear mixed-effects model (LMM), where we incorporated the same fixed effects but included participants as a random-intercepts effect to control for individual differences. The rates of log-transformed antibody titer decay along with the limit of detection of our assay were used to estimate the number of days that the antibodies remain detectable after both natural infection and full vaccination.

Ten-fold cross-validated least absolute shrinkage and selection operator (LASSO) models were employed as a feature-selection and regularization technique. The LASSO models were tuned to select the simplest model within one standard error of the lowest value root-mean-square error accuracy metric that included at least two predictors (Supplementary Table 1). Four LASSO models with identical demographic variables were constructed while controlling for time or days since LPT, 1st dose, or 2nd dose, respectively, including: 1) the effect of infection symptoms on the antibody response post-infection, 2) the effect of infection symptoms on the antibody response post full vaccination, 3) the effect of dose 1 vaccine symptoms on the antibody response post full vaccination, and 4) the effect of dose 2 vaccine symptoms on the antibody response post full vaccination. The selected predictors from each of the best-fitting cross-validated LASSO models were then included as fixed effects in follow-up LMMs with by-participant random intercepts, allowing us to control for individual differences. For significant categorical fixed effects from the LMMs, we conducted post-hoc Tukey tests to confirm directionality and to correct for multiple comparisons.

Additional linear regressions were used to investigate effects of each symptom following infection, 1^st^ dose of vaccination, and 2^nd^ dose of vaccination and explore possible relationships between demographics factors on peak antibody titer levels following full vaccination. Analyses were performed using R statistical software Version 4.1.1 [24]. Generalized additive modeling, cross-validation, LASSO modelling, LMM, and post-hoc Tukey tests were conducted with the R packages *mgcv* [25], *caret* [26], *glmnet* [27], *nlme [28]*, and *glht* [29], respectively, while the linear modelling, Mann-Whitney U tests, and Kruskal-Wallace tests were performed using the R package *stats* [24]. Plots were produced using the *ggplot2* [30].

## Results

### Characteristics of the Study Population

Demographic characteristics are detailed in Table 1. Thirty-three participants were included in our post infection cohort. The median number of days since LPT to entry into the study was 101 days. For the post dose 1 and post dose 2 analysis, we included 49 and 48 participants, respectively.

**Table 1:** Characteristics of the Study Population Following Natural Infection, 1st Dose of Vaccine, and 2nd Dose of Vaccine

|  | Post Infection | Post Dose 1 | Post Dose 2 |
| --- | --- | --- | --- |
| **n** | 33 | 49 | 48 |
| **Gender, M/F** | 14/19 | 17/32 | 16/32 |
| **Race** | White [25 (76%)]<br>Black/African American<br>[2 (6%)]<br>Asian [1 (3%)]<br>Other [5 (15%)] | White [39 (80%)]<br>Black/African<br>American [2 (4%)]<br>Asian [2 (4%)]<br>Other [6 (12%)] | White [(39 (81%)]<br>Black/African American<br>[2 (4%)]<br>Asian [2 (4%)]<br>Other [5 (110%)] |
| **Ethnicity, Hispanic/Not**<br>**Hispanic** | 14/19 (42%/58%) | 22/27 (45%/55%) | 21/27 (44%/56%) |
| **Median age [Range]** | 39 [20-76] | 39 [20-78] | 39 [20-78] |
| **Vaccine manufacturer,**<br>**Pfizer/Moderna** | - | 31/18 (63%/37%) | 30/18 (62.5%/37.5%) |
| **Median days since COVID**<br>**diagnosis by PCR to entry**<br>**SD**<br>**Range** | 101<br>74.87<br>8 - 292 | - | - |
| **Median days since COVID**<br>**diagnosis by PCR to**<br>**vaccination**<br>**95% CI** | - | 99<br>72 – 159 | 127<br>102-180 |

<sup>a</sup> Each count is the number of individuals who self-reported each symptom at the timepoint listed in each column. Percentages are based on the total n for each column.
<sup>b</sup> All samples collected following natural infection were included for analysis.

**Table 1:** Symptoms Experienced by the Study Cohort Following Natural Infection, 1st Dose of Vaccine, and 2nd Dose of Vaccine

|  | Post Infection | Post Dose 1 | Post Dose 2 |
| --- | --- | --- | --- |
| **Symptoms Reported During SARS-CoV-2 Infection<sup>a</sup>** |  |  |  |
| Asymptomatic | 10 (20%) | - | - |
| Anosmia | 27 (55%) | - | - |
| Congestion/rhinorrhea | 26 (53%) | - | - |
| Cough | 23 (47%) | - | - |
| Difficulty breathing | 12 (24%) | - | - |
| Dysgeusia | 26 (53%) | - | - |
| Fatigue | 31 (63%) | - | - |
| Fever | 18 (37%) | - | - |
| Myalgias | 28 (57%) | - | - |
| Nausea or vomiting | 5 (10%) | - | - |
| Sore Throat | 15 (31%) | - | - |
| Upset Stomach | 14 (29%) | - | - |
| **Local Symptoms Reported Following Vaccination<sup>a</sup>** |  |  |  |
| Injection site pain | - | 25 (51%) | 26 (53%) |
| Injection site redness | - | 2 (4%) | 4 (8%) |
| Injection site swelling | - | 4 (8%) | 5 (10%) |
| **Systemic Symptoms Reported** |  |  |  |

| Following Vaccination<sup>a</sup> |  |  |  |
| --- | --- | --- | --- |
| Asymptomatic | - | 22 (45%) | 20 (41%) |
| Chills | - | 9 (18%) | 11 (22%) |
| Fatigue | - | 14 (29%) | 19 (39%) |
| Fever | - | 10 (20%) | 10 (20%) |
| Headache | - | 14 (29%) | 12 (24%) |
| Myalgias/Muscle<br>Aches | - | 12 (24%) | 14 (29%) |

<sup>a</sup> All symptoms reported during SARS-CoV-2 Infection, as well as local and systemic symptoms following vaccination were included in LASSO modeling as shown in Supplementary Table 1.

**Table 1:**
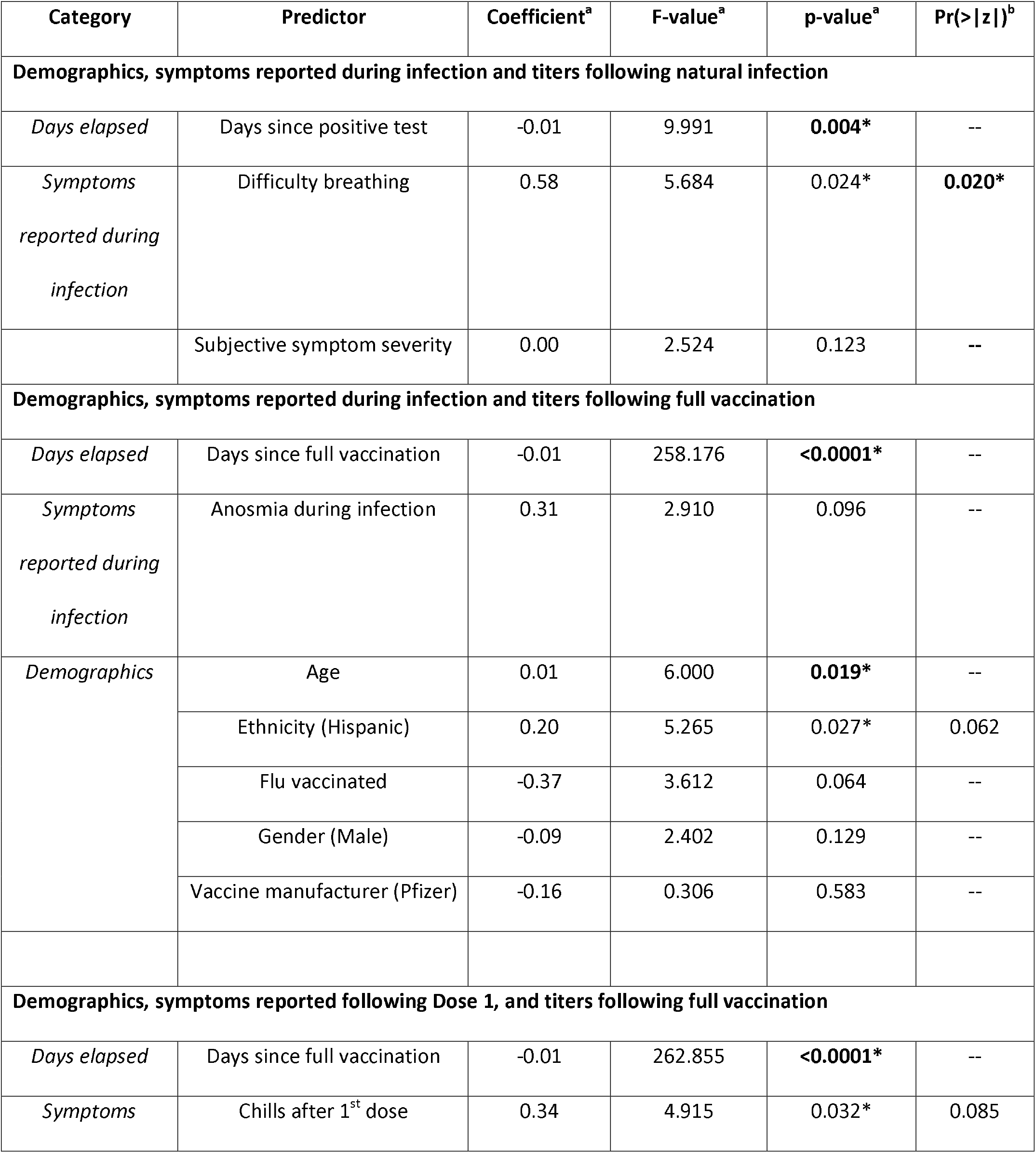
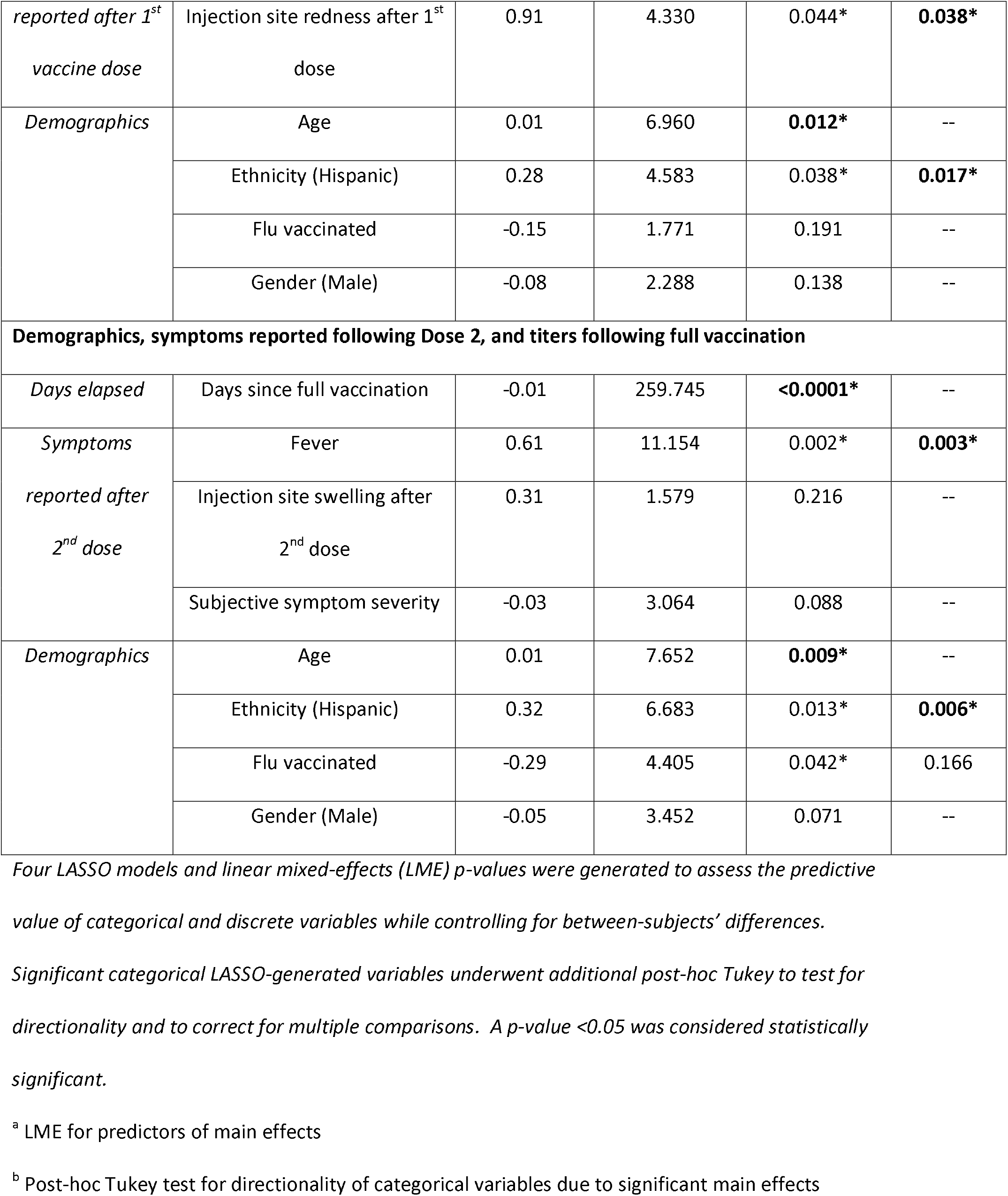
Symptoms and demographic factors influence antibody responses following natural infection and full vaccination

### Symptoms Reported Following Infection and Vaccination

The highest reported symptom during the vaccine-naïve COVID-19 course was fatigue (63%) (Table 2). Other highly reported symptoms included anosmia (55%), congestion (53%), and myalgias/muscle aches (57%). Following dose 1, the most common symptoms were injection site pain (51%), headache (29%), and fatigue (29%). Similarly, the most common symptoms reported following the second mRNA vaccination (dose 2) were injection site pain (53%), fatigue (39%), and myalgias (29%) (Table 2).

### In Previously Infected Individuals, Antibody Titers Are More Robust Following Full-Vaccination as Compared to Post Natural Infection

We found that there was a more robust antibody response immediately following full vaccination (Figure 1C) when compared to the antibody response following natural infection (estimate = 4.117, *t* = 12.950, *p* = < 0.001) (Figure 1B), where peak log_2_ antibody titers where greater in the vaccination with prior infection (14.517) than in natural infection (9.217). This is illustrated in Figure 1A, where natural infection and post-full vaccination titers were included in a bi-phasic model to show longitudinal antibody responses. Infected individuals had a slower rate of antibody titer decay (−0.010 vs -0.015 log per day), though this effect was small (estimate = -0.005, *t* = 2.351, *p* = 0.020). Our linear model also predicted that antibody titers remain detectable for a longer period of time after full vaccination (535 days) compared to natural infection alone (404 days), using the respective rates of antibody decay and the limit of detection for our assay. Additional LMMs confirmed our findings that antibody titer declined faster following full vaccination than in natural infection (estimate = -0.006, *F* =11.238, *p* <0.001). Of note, the combination of natural infection followed by vaccination, or so-called “hybrid immunity”, elicits a more durable antibody response than natural infection alone (estimate = 4.138, *F* = 794.623, *p* < 0.0001), as log_2_ antibody titers were predicted to remain detectable for a longer period of time following vaccination (550 days) than natural infection (464 days).

**Figure 1:**
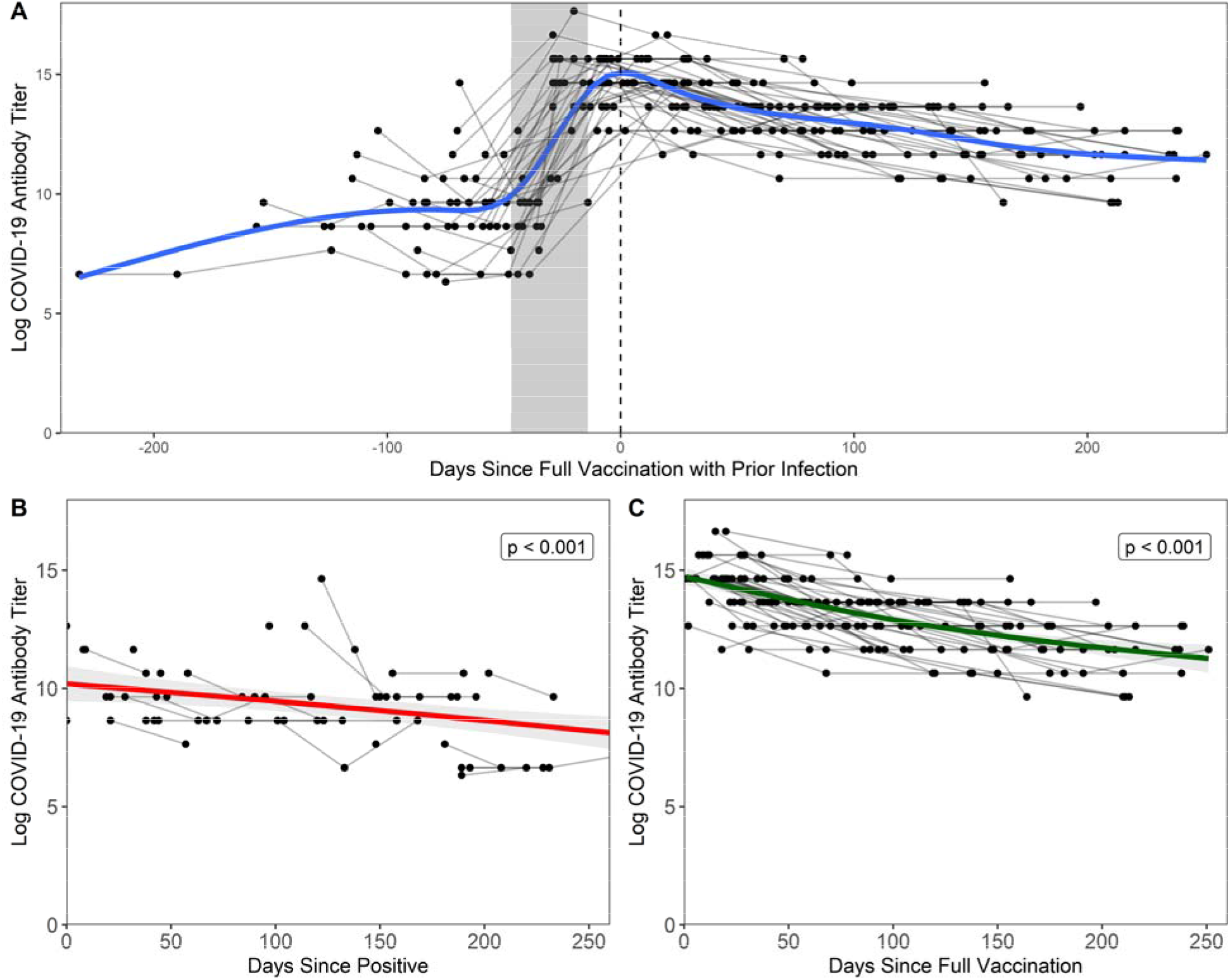
Antibody response following natural infection and vaccination. Each black point represents a sample from a participant, grey lines connect points from the same participant, and the grey shaded area represents the maximum number of days between doses relative to date of full vaccination (14 days after second dose, regardless of vaccine manufacturer). A) Days since full vaccination vs. log titers over time. t=0 on the x-axis represents the day when COVID+ participants became fully vaccinated (2 weeks after second vaccination). Bi-phasic, generalized additive model (GAM) is visualized by a blue line. B) In unvaccinated COVID+ participants, log_2_ antibody titers decay at a rate of -0.010 per day after last positive COVID-19 test result. Fitted linear model is visualized by a red line. Note that three points were excluded from the above figure due to the temporal scale used to graphically depict the data but are included in the analyses herein. C) In vaccinated COVID+ participants, log_2_ antibody titers decay at a rate of -0.015 per day after full vaccination. Fitted linear model is visualized by a green line.

**Figure 1:**
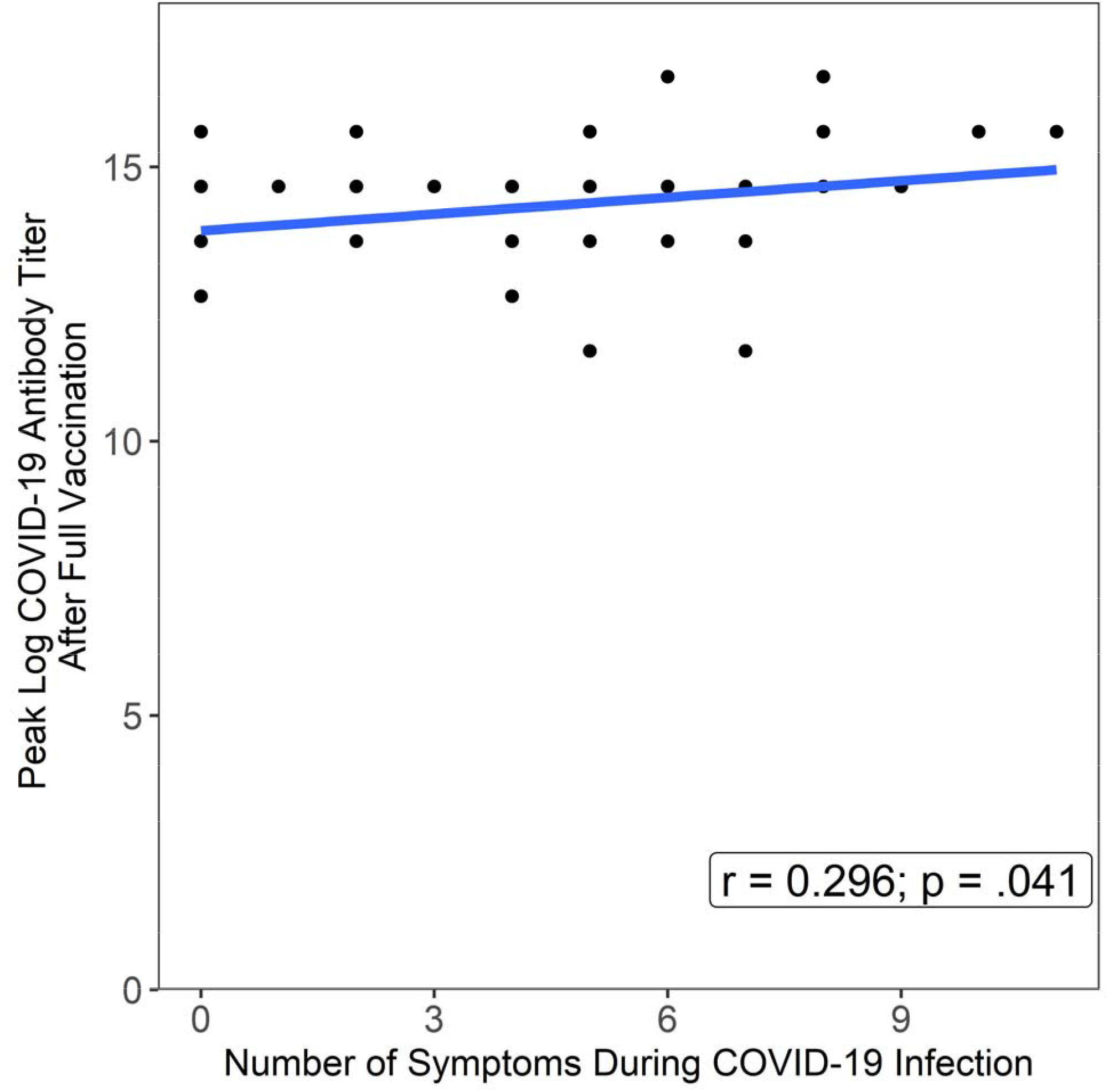
Symptoms experienced during infection significantly influences peak titer responses following full vaccination. Number of symptoms during COVID-19 infection were positively correlated with peak log_2_ COVID-19 antibody titers following full vaccination (Pearson’s r = 0.296; p = .041)

### Days Elapsed and Symptoms Reported During Infection Influence Antibody Titers

As shown in Table 3, difficulty breathing during infection (estimate = 1.590, *F* = 5.684, *p* = 0.024) and days elapsed since LPT (estimate = -0.006, F = 9.912, *p* = .004) were significant main effects in predicting antibody titers following natural infection. Post-hoc testing confirmed that antibody titers were elevated in individuals who experienced difficulty breathing (*z* = 2.612; *p* = 0.009).

When modeling demographics and symptoms at infection to predict the antibody response after full vaccination, we found that days elapsed since full vaccination (estimate = -0.014; *F* = 258.176; *p* < 0.0001), age (estimate = 0.018; *F* = 6.000; *p* = 0.019), and ethnicity (Hispanic) (estimate = 0.456; *F* = 5.265; *p* = 0.018) were significant main effects, though no categorical variables were significant after post-hoc testing.

### Symptoms Following Vaccination Are Predictive of Higher Antibody Titers After Full Vaccination

Local and systemic symptoms following 1^st^ dose of SARS-CoV-2 mRNA vaccines were predictive of higher antibody titers after full vaccination. As seen in Table 3, the results of the LMM show that days elapsed since full vaccination (estimate = -0.014, *F* = 262.855, *p* < 0.0001), chills (estimate = 0.541; *F* = 4.915; *p* = 0.032), injection site redness (estimate = 1.243; *F* = 4.330; p = 0.044), age (estimate = 0.021; *F* = 6.960; *p* = 0.012), and ethnicity (estimate = 0.562, *F* = 4.583, *p* = 0.038) were significant. Following post-hoc Tukey testing on the significant categorical main effects, we found that injection site redness (*z* = 2.081, *p* = 0.038) and ethnicity (Hispanic) (*z* = 2.382, *p* = 0.017) were significant, while the main effect of chills was not.

We also examined demographics and symptoms reported during the 2^nd^ dose of SARS-CoV-2 mRNA vaccination and their effect on the antibody response after full vaccination. Days since full vaccination (estimate = -0.014; *F* = 259.745; *p* < 0.0001), age (estimate = 0.023; *F* = 7.652; *p* = 0.009), identifying as Hispanic (estimate = 0.609; *F* = 6.683; *p* = 0.013), fever (estimate = 0.839; *F* = 11.154, *p* = 0.002), and influenza vaccination (estimate = -0.475; *F* = 4.405; *p* = 0.042) were observed to be significant. Fever and ethnicity (Hispanic) were found to be statistically significant (*z* = 3.016, *p* = 0.003; *z* = 2.735, *p* = 0.006, respectively) following post-hoc testing, though influenza vaccination was not.

### The Number of Symptoms During Infection Result in Higher Peak Antibody Titers Post Full-Vaccination

As shown in Figure 2, the number of symptoms reported during infection significantly predicted peak antibody titers after full vaccination (estimate = 0.10, *t* = 2.10, Pearson’s *r* = 0.296; *p* = .041). Additional linear models were conducted for the number of symptoms reported as a function of demographics, where we found that the number of symptoms self-reported during infection was significantly influenced by self-identifying as White (estimate = 4.679, *t* = 2.153, *p* = 0.037). No other demographic was significant.

## Discussion

The goal of this study was to investigate the role of demographics, pre-existing immunity, and symptomatology following infection and vaccination to ascertain whether they independently or collectively are associated with immunogenicity following mRNA vaccination for COVID-19 in individuals previously infected with SARS-CoV-2. Our results demonstrate higher durability and robustness of antibody titers despite a faster rate of antibody decay following vaccination, which supports previously reported findings [31] for SARS-CoV-2 mRNA vaccines. Unsurprisingly, our results also demonstrate that a larger temporal gap between an individual’s LPT and/or date of vaccination predict decline of antibody titers over time.

Following infection alone, we found that the number of symptoms reported and difficulty breathing during the COVID-19 course were predictive of higher antibody titers. This result supports existing evidence [32, 33] that individuals who report a more severe or symptomatic SARS-CoV-2 infection have higher peak titers than asymptomatic individuals. Our data also support higher peak antibody titers following vaccination with increasing age [32, 34]. The possible relationship between the antibody titer post-vaccination and vaccine reactogenicity is largely unknown. Given that strength of the antibody response was shown to correlate with disease severity in patients with COVID-19, we hypothesized that more prominent post-vaccination adverse reactions might be associated with a stronger immune response. After receiving dose 1 of either Pfizer or Moderna’s COVID-19 vaccine, the local symptom of injection site redness was found to be significantly predictive for higher antibody titers following full vaccination and could be an indicator of the early and more prominent immune response. Interestingly, after dose 2 we found that fever was significantly predictive for higher antibody titers following full vaccination, though it should be noted that asymptomatic individuals mounted robust immune responses as well.

Most critically, our analysis demonstrates that in previously infected individuals, SARS-CoV-2 mRNA vaccines result in a more robust antibody response than that following infection alone. Indeed, in individuals with “hybrid immunity”, antibody titers peak following full vaccination at 4-fold higher than those following naturally acquired immunity and appear to persist at detectable levels for >500 days following vaccination. One explanation for this increased response could be the presence of pre-existing memory T and B cell responses developed during natural infection. These cells might enhance a secondary immune response following vaccination similar to that of a booster immunization. In addition to bolstering the current CDC recommendations [35] that individuals previously infected with SARS-CoV-2 receive vaccination, our results provide additional, longitudinal support for this measure. In addition to lowering the risk of re-infection [36, 37], vaccination clearly increases an individual’s immune response [38, 39], which is vitally important as SARS-CoV-2 continues to produce variants capable of immune evasion, such as Delta (B.1.617.2) and Omicron (B.1.1.529) [40].

Current evidence supports a stronger reactogenic profile (both systemic and local) following the first vaccine dose in previously infected individuals [41, 42], though it is worth noting that the median time between COVID-19 to the first dose of vaccine in the studies cited above was either not reported or was only 2.9 months. In our cohort, there is considerable variability between participants in days elapsed from LPT to time of vaccination (1^st^ dose median = 99 days; 95% CI = 72–159 days; 2^nd^ dose median = 127 Days; 95% CI = 93-180 days). Given current knowledge regarding antibody decline, it is reasonable to posit that a longer temporal gap between SARS-CoV-2 infection and vaccination may have resulted in a more reactogenic response to the 2^nd^ dose of vaccine when compared to participants who had shorter intervals between infection and vaccination as noted in the studies described above.

Previous work [43] examining the relationship between SARS-CoV-2 vaccine reactogenicity and immunogenicity has been limited by assay type (i.e., semi-quantitative assays) and only deigned to measure immunity immediately following vaccination. Another group [44] conducted a survey to assess reactogenicity of COVID-19 vaccines and found that previous infection was associated with an increased risk of reporting any side effect. They also found that mRNA vaccines seemed to produce milder, less frequent systemic side effects but more local reactions in comparison to vector vaccines (i.e., Oxford AstraZeneca AZD1222), which are consistent with our findings following the 1^st^ dose. To our knowledge, we are the first to examine predictors of reactogenicity and immunogenicity following SARS-CoV-2 vaccines in participants with prior COVID-19.

Intrinsic factors, such as age, gender, and ethnicity are thought to influence immunogenicity, but little is known about the impact of ethnicity in particular [19]. Our study was conducted in Miami-Dade County, an international, multi-cultural hub where the population is largely Hispanic and bilingual. Notably, our analysis demonstrated a significant relationship between ethnicity (Hispanic) and higher antibody titers over time at nearly every time point of interest, including infection where the analysis approached significance (*p* = 0.0624). Other groups have demonstrated higher rates of Hispanic SARS-CoV-2 seroconversion when compared to other ethnicities [45, 46] and have found that Hispanic ethnicity is linked to higher rates of seroprotection and seroconversion following H1N1 monovalent vaccination [47, 48], but additional future studies with a large number of participants are needed to support a generalizable trend for antibody magnitude over time in this population.

In addition, we found that influenza vaccination was associated with higher antibody titers in our model examining symptoms following the 2^nd^ dose and antibody titers following full vaccination, though it was not found to be significant following post-hoc testing. Although biological relevance for this finding is unknown, our group has previously shown that specific A(H1N1) CD4 responses correlate with SARS-CoV-2 specific CD4 T-cells, suggesting a protective effect of pre-existing influenza specific T-cells [7]. We speculate that this finding provides evidence for healthy and “trained” immune systems within our cohort, wherein epigenetic and metabolic reprogramming have augmented innate immune cells that enhance adaptive immunity to increase SARS-CoV-2 specific responses [7, 49].

Our study has several limitations. Sample sizes for each cohort examined were small due to variability in vaccination timelines and participant scheduling. Some individuals were excluded due to a confounding effect on our predictive modeling, which is controlled by the fixed effect of time. The natural infection group was further limited by the study timeline, as the first SARS-CoV-2 vaccination became available shortly after enrollment began and therefore limited the number of individuals we were able to follow longitudinally. Additionally, our analysis only included quantitative antibody binding titers. Although recent work has demonstrated that higher binding antibodies correlate to higher neutralizing antibodies [13], expansive, multi-center longitudinal studies are needed. An ideal analysis would consist of a multivariate analysis of reactogenicity, demographics, and quantitatively characterized antibody, B-cell and T-cell responses, as immune protection seems to be contingent on all three tiers of the immune response [50]. Further, some of the predictors used in our statistical analysis were found to be significant in one test but not in post-hoc tests. Large, longitudinal studies are required to confirm a significant group difference, but the predictors utilized herein should be included in future analyses. Our bivariate analysis of symptoms experienced following the 1^st^ and 2^nd^ doses failed to demonstrate that individual symptoms can influence peak antibody titers following full vaccination. The same was true for race and ethnicity, which were not found to be significantly predictive for peak titers over time, though we contend that this is because these models failed to control for individual differences, or intercepts, to account for between-subjects’ variability.

In conclusion, this work supports vaccination in COVID-19+ individuals to assure the most robust immune response possible. A combination of systemic and local symptoms is predictive of higher antibody titers, which may correlate to a higher degree of protection. While more work is needed to understand the role of antibodies against SARS-CoV-2 infection and breakthrough infection, particularly in the age of boosters and variants capable of immune escape, repeating this type of analysis at the population level will be critical in providing individual recommendations for future vaccine measures.

## Data Availability

All data produced in the present study are available upon reasonable request to the authors.

## Acknowledgements

We express our sincerest gratitude to our intellectually curious and willing study participants for donating their blood and time to this study. We also thank the core research team that have made this study logistically possible, including Margaret Roach, Elizabeth Varghese, Celeste Sanchez, Ailet Reyes, and the entire team of phlebotomists at the University of Miami’s Clinical Translational Research Site.

