## Supplementary Table 1 for "Predictors for Reactogenicity and Humoral Immunity to SARS-CoV-2 Following Infection and mRNA Vaccination: A Regularized Mixed-Effects Modelling Approach"

Supplementary Table 1: Demographics and symptoms included in LASSO modelling

|  | **Included in LASSO Following Natural Infection** | **Included in LASSO Following 1^st^ Dose** | **Included in LASSO Following 2^nd^ Dose** |
| --- | --- | --- | --- |
| **Demographics** |  |  |  |
| Age**^a^** | X | X | X |
| Days Elapsed Since Positive Test**^a^** | X |  |  |
| Days Elapsed Since Full Vaccination**^a^** |  | X | X |
| Ethnicity [Hispanic/Not Hispanic] | X | X | X |
| Flu Vaccination Status | X | X | X |
| Gender [Male/Female] | X | X | X |
| Race [Black/White/Other] | X | X | X |
| Vaccine Manufacturer [Pfizer/Moderna] | X | X | X |
| **Symptoms Following Infection or Vaccination** |  |  |  |
| Anosmia | X |  |  |
| Chills |  | X | X |
| Congestion/Rhinorrhea | X |  |  |
| Cough | X |  |  |
| Difficulty Breathing | X |  |  |
| Dysgeusia | X |  |  |
| Fatigue | X | X | X |
| Fever | X | X | X |
| Headache |  | X | X |
| Injection Site Pain |  | X | X |
| Injection Site Redness |  | X | X |
| Injection Site Swelling |  | X | X |
| Myalgias | X | X | X |
| Nausea/Vomiting | X |  |  |
| Sore Throat | X |  |  |
| Symptom Severity**^a^** | X | X | X |
| Upset Stomach | X |  |  |

Categorical and discrete variables were included for generation of LASSO following natural infection, dose 1, and dose 2. Variables that were found to be significant were included in subsequent LMM.

^a^ non-categorical (discrete) variable
